# Age-based differences in quantity and frequency of consumption when screening for harmful alcohol use

**DOI:** 10.1101/2022.03.15.22272307

**Authors:** Sarah Callinan, Michael Livingston, Paul Dietze, Gerhard Gmel, Robin Room

## Abstract

**Aims:** Survey questions on usual quantity and frequency of alcohol consumption are regularly used in screening tools to identify drinkers requiring intervention. The aim of this study is to examine age-based differences in quantity and frequency of alcohol consumption on the Alcohol Use Disorders Identification Test (AUDIT) and how this relates to the prediction of harmful or dependent drinking

**Design:** Cross sectional survey

**Setting:** Australia.

**Participants:** Data was taken from 17,399 respondents who reported any alcohol consumption in the last year and were aged 18 and over from the 2016 National Drug Strategy Household Survey, a broadly representative cross-sectional survey on substance use.

**Measurement:** Respondents were asked about their frequency of consumption, usual quantity per occasion and the other items of the AUDIT.

**Findings:** In older drinkers, quantity per occasion (β=0.53 (0.43, 0.64 95%CI in 43-47 year olds as an example) is a stronger predictor of dependence than frequency per occasion (β=0.24 (0.17, 0.31). In younger drinkers the reverse was true with frequency a stronger predictor (β=0.54 (0.39, 0.69) in 23-27 year olds) than quantity (β=0.26 (0.18, 0.34) in 23-27 year olds). Frequency of consumption was not a significant predictor of dependence in respondents aged 73 and over (β=-0.03 (−0.08, 0.02)). Similar patterns were found when predicting harmful drinking. Despite this, since frequency of consumption increased steadily with age, the question on frequency was responsible for at least 65% of AUDIT scores in drinkers aged 53 and over.

**Conclusions:** The items with a weaker association with dependent or harmful drinking in younger and older drinkers are the same items with the strongest influence on overall AUDIT scores. Further investigation into age-specific scoring of screening tools is recommended.

## INTRODUCTION

Alcohol is responsible for 2% and 7% of death and disability-adjusted life years lost in women and men respectively (1), and this burden falls substantially on people with an alcohol use disorder (2). Reliable identification of those who may benefit from appropriate interventions is a vital part of reducing alcohol related harms (3). Identification of possible alcohol use disorders often relies on screening tools that can be administered by clinicians or other health professionals. One of the most commonly used screening tools for harmful drinking and/or possible alcohol use disorder is Alcohol Use Disorders Identification Test (4, 5), which is used around the world after being translated into at least 46 languages (6).

The AUDIT was written to assess three constructs related to alcohol use disorders; three items on consumption, three that make up a dependence subscale and four that address adverse consequences of harmful drinking (7). There is evidence to suggest that the overall AUDIT score is a good screening tool for dependence (8) and that it can identify alcohol use disorders with excellent concurrent validity (9). Much of the power of the AUDIT is derived from the first three questions on consumption, typically referred to as the AUDIT-C. This subscale asks about the frequency of drinking, the quantity consumed per usual occasion and the frequency of risky drinking occasions in the past year. A high proportion of the average AUDIT score comes from the AUDIT-C; 90% of the score from the ten item AUDIT in a Swedish study came from the AUDIT-C; this dropped to 71% if you excluded anyone with a full AUDIT score of less than eight (10). Interestingly, Selin (2006) found that the correlation between the AUDIT-C and the subscale on harmful drinking was only *r* = 0.49, indicating that these measures of consumption are not always the best indicator of harmful drinking. Another study found that the frequency item in particular was a poor indicator of harmful drinking or dependence (11).

Frequency per occasion has been found to rise with age while quantity per occasion decreases (12). Despite these differences in drinking patterns, the same screening tools are used for younger and older drinkers. This may have implications for the efficacy of these tools at identifying genuinely problematic drinking. Heavier drinking occasions are more common in younger drinkers (12), meaning their usual quantity per drinking occasion tends to be higher. For example, many young people who drink heavily in their youth cease this practice without needing treatment as they age, this phenomenon has been referred to as “transient teenage drinking” (13), or a “teen transient false-positive” (14) in those less than 20, or “spontaneous remission” (15) in those under 40,. Given this trend, it’ s possible that a high usual quantity per occasion is not as clearly indicative of a need for intervention in young people the same way it might for an older drinker. If this is true, it is also possible that there might be other measures that could more accurately identify those drinkers who do not age out of these harmful drinking patterns and would thus benefit from intervention.

There has been little research assessing validity of the AUDIT across age groups with widely varying patterns of drinking. By using the dependence and harmful drinking subscales of the AUDIT, it is possible to assess how quantity and frequency of consumption are related to dependent and/or harmful drinking and whether this differs by age. This exploratory study will do the following:

1. Estimate how average drinking frequency and quantity per drinking occasion vary by age
2. Test the predictive validity of average drinking frequency and usual drinking quantity in predicting dependence as measured by the AUDIT in age stratified groups.
3. Test the predictive validity of these constructs in predicting harmful drinking as measured by these two subscales of the AUDIT in age stratified groups.
4. Calculate the relative contribution from the first two questions on quantity and frequency to full AUDIT scores in each age group.

## METHODS

### Design

Cross sectional correlational design using data from a survey broadly representative of the Australian population. Analyses were not pre-registered and should thus be considered exploratory. This work was done in line with a STROBE checklist, found in Supplementary Table 1.

### Materials and sample

The Australian National Drug Strategy Household Survey (NDSHS) is a national, cross-sectional survey of alcohol and drug use patterns, attitudes and behaviours conducted every three years. The current study uses data from the 2016 wave (16), the most recent wave with all the questions from the AUDIT included. The survey was administered to a geographically-stratified random sample of 23,772 English speaking Australians aged 14 years and above living in private dwellings. In selected households with more than one eligible respondent, the person with the most recent birthday was selected, with a response rate of 51.1% (16). Respondents completed the surveys via either a paper form which was administered via a drop-and-collect method, an online form or a computer-assisted telephone interview (16). Respondents aged under 18 (legal drinking age in Australia) were excluded from the current study, leaving 22,642 respondents. Respondents who reported consuming no alcohol in the past twelve months were also excluded, leaving 18,520 respondents. Finally, 6.05% of the sample did not answer the questions on our two main variables of interest, usual quantity per occasion and frequency of drinking occasions. Both harmful drinking (t(18062)=4.84, *p* < .001) and dependence (t(18116)=3.62, *p* < .001) sub-scores were lower in those who were missing on this item compared those who were included in the study (among those who also answered the questions on each subscale respectively), indicating that the remaining sample may have a slight overrepresentation of heavier drinkers. These respondents were excluded, leaving a final sample of 17,399 respondents (52.7% female) with a mean age of 49.96 (SD = 17.39). Please note that there are also some respondents who are removed from each analysis including an outcome variable on which they were missing; the dependence subscale (0.80%, remaining N = 17260), harmful drinking subscale (1.06%, remaining N = 17215) or the full AUDIT (7.46%, remaining N = 16101). However, they have not been removed from analyses on which they have no missing data – i.e., if a respondent is missing on the dependence subscale but not the harmful drinking subscale, they are included in the analysis on harmful drinking. As above, those who were missing on these outcome variables appeared to be lighter drinkers than those in our final sample (t(17397)=16.12, *p*<.001) based on their total volume of consumption (quantity x frequency). A figure outlining the eligible and missing data in the study can be found in Supplementary Figure 1.

### Measures

#### Predictor variables

##### Frequency of consumption

Respondents are asked how often they have consumed alcohol, with multiple response options. The mid-point of each response option (i.e., 2-3 days per week = 2.5 days per week) was used to calculate the number of drinking occasions per year. This was then divided by 52 to get drinking occasions per week. This was used for Figures 1 to 3. The conversion of these response options to match those in the first question of the AUDIT for Figure 4 is shown in Supplementary Table 2.

**Figure 1.**
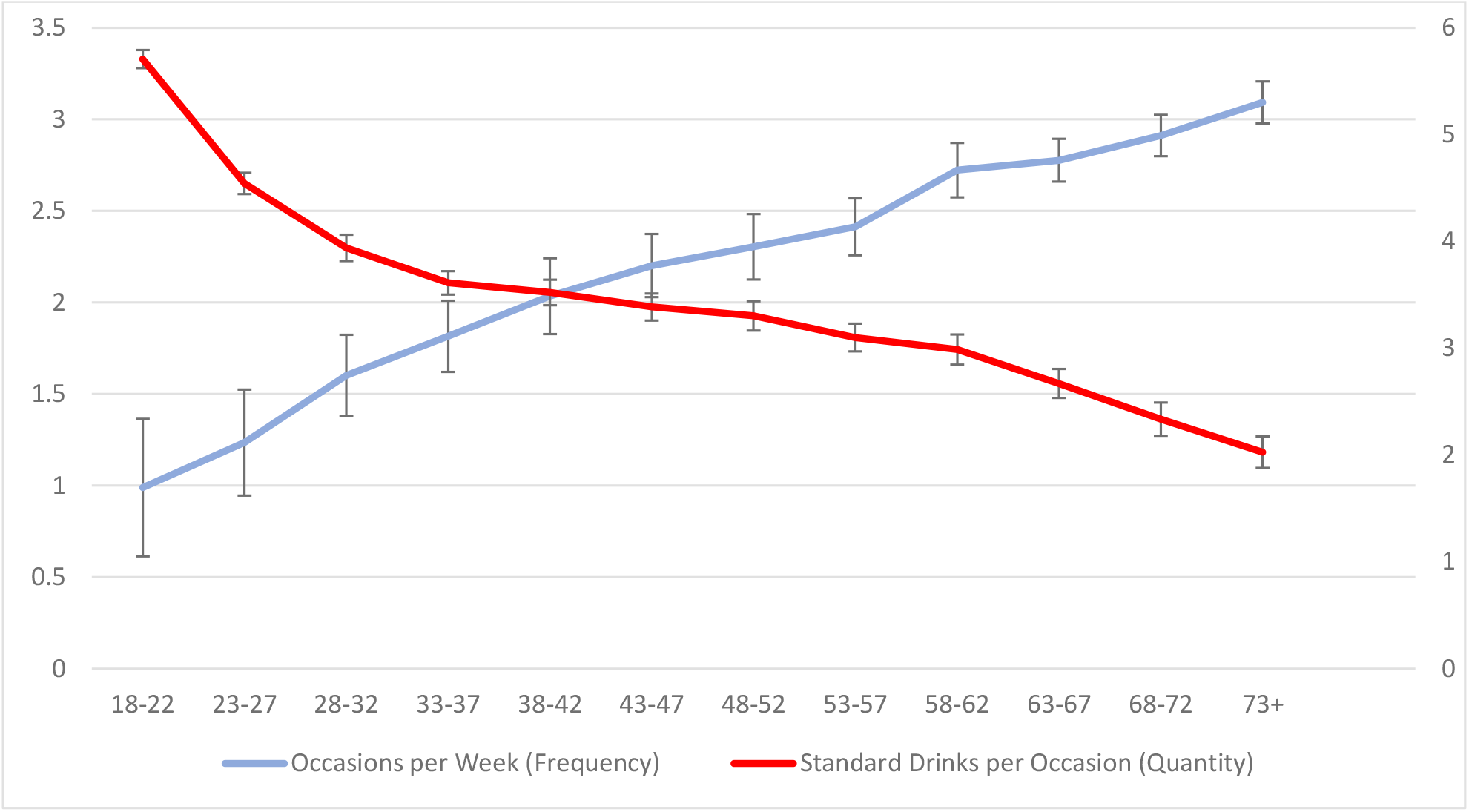
Average number of drinking occasions per week (frequency, left scale) and number of standard drinks per usual occasion (quantity, right scale), with 95% confidence intervals, by age N=17,399

**Figure 2.**
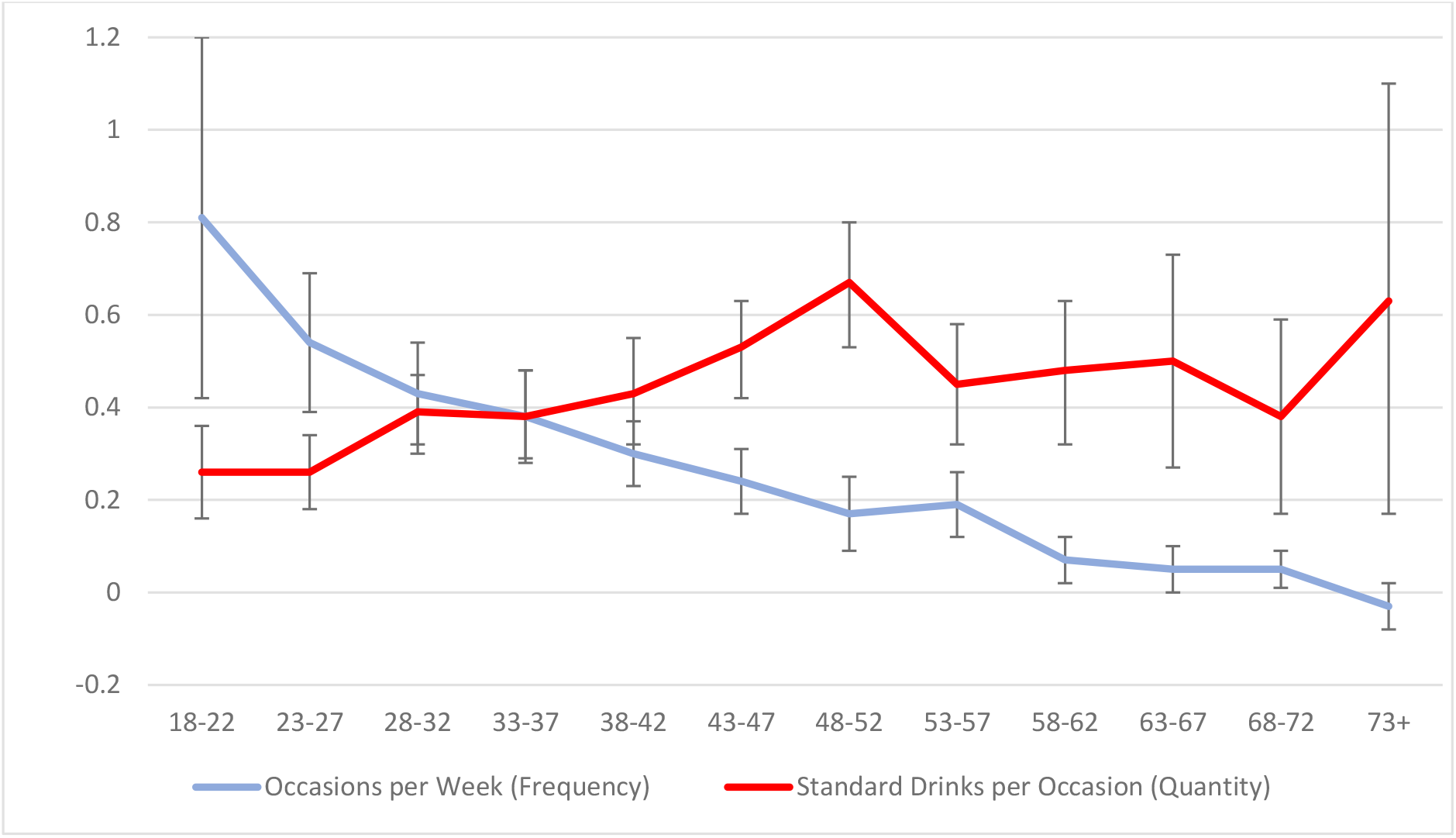
Beta coefficients of standardised usual quantity and frequency of drinking as predictors of the AUDIT dependence subscale. N=17,260

**Figure 3.**
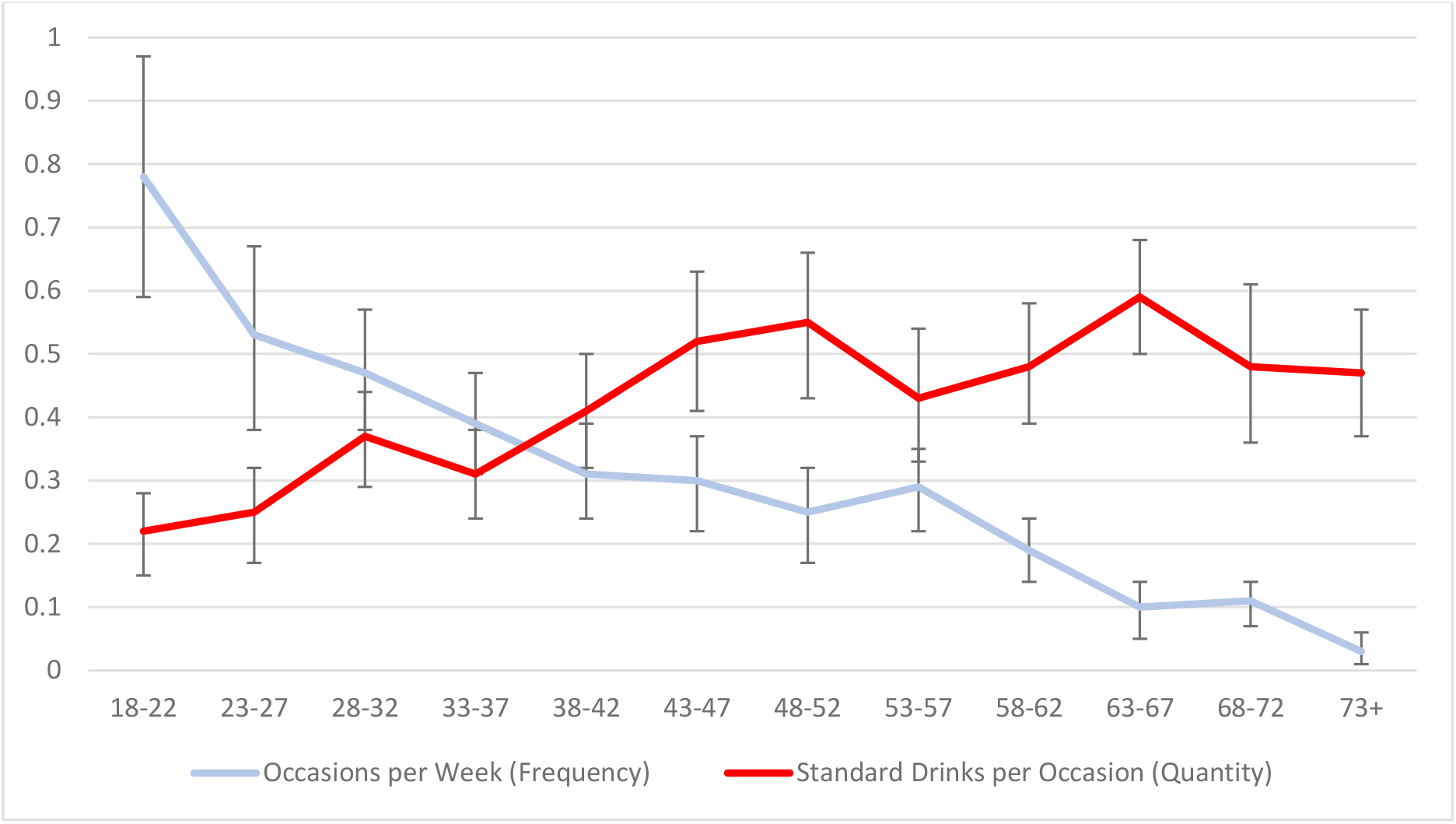
Beta coefficients of standardised usual quantity and frequency of drinking as predictors of the AUDIT harmful drinking subscale. N=17,215

**Figure 4.**
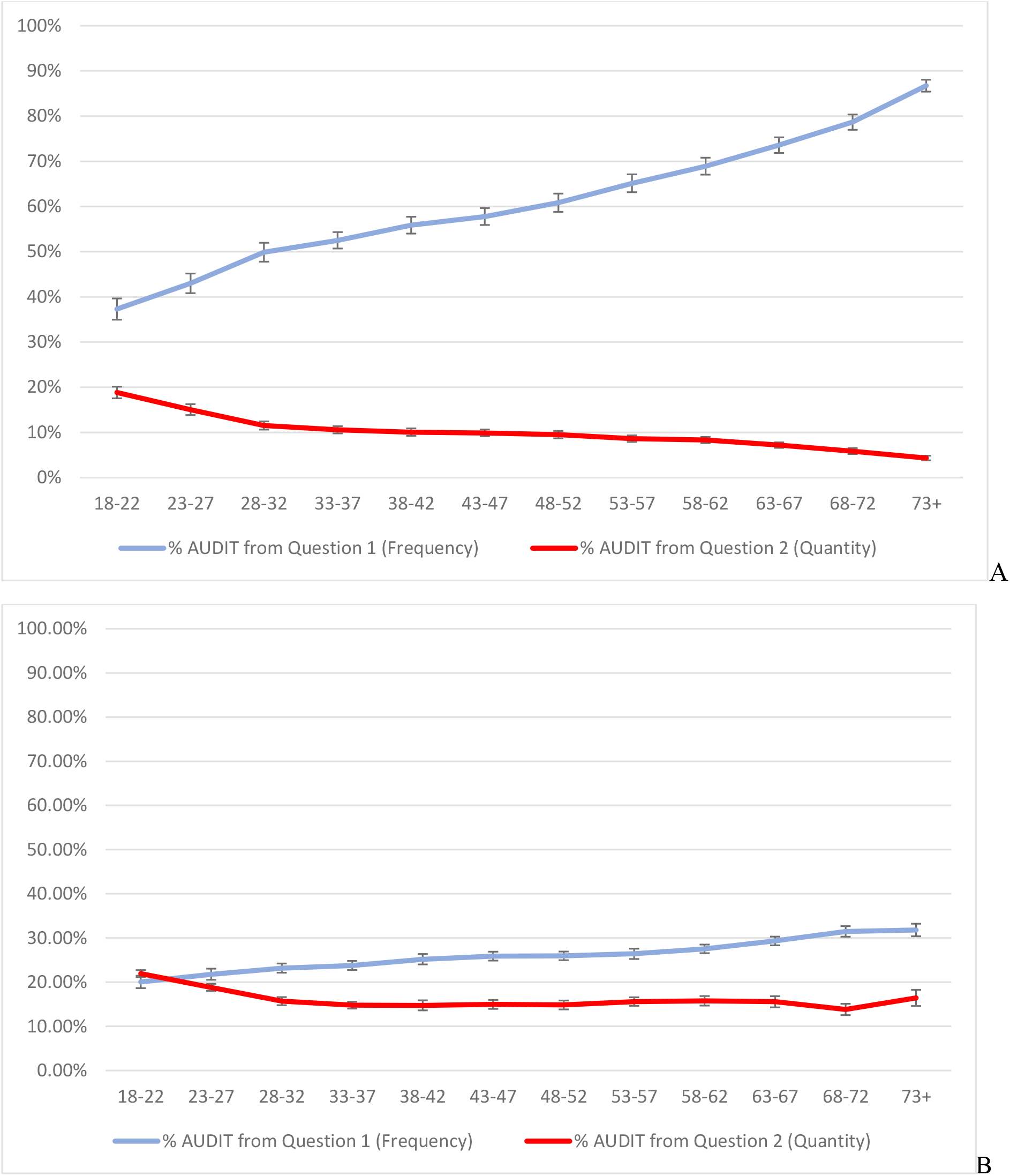
Proportion of the full AUDIT score that can be attributed to the question on frequency (item 1) and usual quantity (item 2). Panel A includes all participants, Panel B includes only those who had a total AUDIT score of 8 or more. N=16,101 (A) and 4,159 (B).

##### Quantity per occasion

This was taken from the item asking “*how many drinks containing alcohol do you have on a typical day when you are drinking?”*. The response is measured in standard drinks (10g ethanol). This was used for Figures 1 to 3. The conversion of these response options to match those in the second question of the AUDIT for Figure 4 is shown in Supplementary Table 2.

#### Outcome variables

##### AUDIT

The Alcohol Use Disorders Identification Test (4) is a ten item scale with a score range of 0 to 40. The items are not administered as a distinct scale in the NDSHS, but all items are covered sufficiently for use (17). All items have a reference period of one year, except for items 9 and 10 where respondents are asked to differentiate between experiences that occurred in the past year or in their lifetime. Item 1 (frequency) and 2 (usual quantity per occasion) are administered as described above. Item 3, on risky single occasion drinking, also had to be derived from other questions in the NDSHS; this is described in full in Supplementary Table 2. The other seven items from the AUDIT are asked as they are in the original scale (4). The authors of the scale recommended a cut-point of eight when wanting to focus on those who would potentially be identified as being in need of intervention (7) – this cut point is used here for the sensitivity analysis focusing on heavier drinkers in Figure 4. The method used to calculate AUDIT scores from the items in the NDSHS survey, outlined in Supplementary Table 2, is based on that used by O’ Brien and colleagues (17). The Cronbach’ s alpha of the full scale is α = 0.80, indicating good reliability. *Dependence subscale:* The summed total of items 4-6 of the AUDIT, with a possible score range of 0 to 12, make up the dependence subscale. The Cronbach’ s alpha of the full scale is α = 0.67, indicating questionable, but not unacceptable, reliability.

##### Harms subscale

The summed total of items 7-10 of the AUDIT, with a possible score range of 0 to 12, make up the harmful drinking subscale. The Cronbach’ s alpha of the full scale is α = 0.60, indicating questionable, but not unacceptable, reliability.

##### Age

Age was treated as a categorical variable, with respondents grouped into five-year age groups starting at age 18 (18-22, 23-27 etc until 73+). These groups were chosen so that any gradual shifts in our results over age can be observed while still ensuring a large sample size in each age group. The size of each age group varied from 871 to 1,774 respondents in the analysis in Figure 1 while in the sub-sample analysis in Figure 4 the size of each age group was between 147 and 471.

### Analysis

All analyses were completed using Stata version 15 (18). Population weights were used in all analyses to ensure that estimates were as representative of the Australian population as possible (benchmarks were based on sub-populations defined by geography, age group and gender). To address aim 1, the mean quantity and frequency of consumption within each age group was calculated. To address aims 2 and 3, beta coefficients from multiple regression models predicting total subscale scores, stratified by age group, were estimated. To enable comparisons of coefficients within each model, both the quantity and frequency variables were converted to z-scores (mean = 0, SD = 1) before being entered as predictors. The outcome variables, harmful and dependence subscale totals, were also converted to z-scores to enable comparisons between the age stratified models. Finally, the focus up to this point is on how the constructs of quantity and frequency related to AUDIT scores so the more detailed, continuous measures available in the NDSHS were used. For aim 4 the focus shifted to how the items on quantity and frequency worked in the AUDIT, so for this analysis the corresponding item scores for these measures were used. The proportion of full AUDIT scores that came from the questions on quantity and frequency (e.g., the score from question one [frequency] divided by the full AUDIT score) were calculated and stratified by age for Figure 4. Finally, 95% confidence intervals for all means, percentages and beta coefficients were calculated and included in all analyses.

## RESULTS

In order to first establish the need to stratify analyses by age the moderating effect of age on the relationship between the predictor variables (quantity and frequency of consumption) and the outcomes (dependence and harms subscale scores) regression models with age x predictor interactions were run. In all four models the interaction was significant (*p*<.001) – the coefficients from the models can be found in Supplementary Table 3.

### Mean frequency of consumption and average quantity per occasion by age

The mean number of drinking occasions per week and drinks per occasion, stratified by age, are shown in Figure 1. The number of drinks per occasion drops quickly from age 18 to 32 and then continues to drop steadily through the older age groups. Conversely, the number of occasions per week rises steadily with the respondent’ s age.

### Predicting harmful drinking and dependence subscales of the AUDIT with usual quantity and frequency of consumption

Coefficients from multiple regression models predicting the dependence subscale score from the AUDIT with usual quantity and frequency as standardised predictor variables, stratified by age, are shown in Figure 2. Using non-overlapping confidence intervals as an indicator of significantly different beta coefficients, frequency of consumption was a better predictor of dependence subscale scores than usual quantity consumed per occasion in respondents aged 18-27. Conversely, quantity of consumption was a stronger predictor of the dependence subscale scores in respondents aged 43 and over.

Figure 3 shows the results from the same analyses replicated for the harms subscale. Again, frequency of drinking was a better predictor of the harmful drinking subscale of the AUDIT than usual quantity consumed per occasion in respondents aged 27 and younger. In respondents aged 43 and over, with the exception of drinkers aged 53-57, quantity per occasion was a better predictor of harmful drinking than frequency.

Next, full AUDIT scores were calculated and the mean proportion of the total score that could be accounted for by the first question (frequency) and second question (usual quantity) of the scale was calculated – these figures are shown in Figure 4A. As can be seen, the frequency question accounts for a high proportion of the total scale, ranging from 37% in 18-22 year olds to 87% in those aged 73 and over. Conversely, only 4% of the score from the oldest age group came from the question on usual quantity of consumption while 19% of the score of 18-22 year olds was accounted for by this question.

To check if this pattern was being primarily driven by those drinkers who did not score high enough to be considered worthy of potential intervention, the same analysis was run again including only those with an AUDIT score of eight or higher. The proportion of the total score driven by these two questions did drop significantly, however the basic pattern was similar. Older drinkers’ AUDIT scores were still more driven by the question on frequency. Younger drinkers’ scores (aged 18-22) were equally accounted for by both usual quantity and frequency -- around 20-22% each. The proportion accounted for by frequency rose with age, while the proportion accounted for by usual quantity dropped with age.

## DISCUSSION

The aim of this paper was to examine the reported usual quantity and frequency of alcohol consumption by age and how these variables predict harmful or dependent drinking as measured by AUDIT subscales. Overall, we found that young people drink more per occasion, but they drink less often, while older drinkers drink more often, but drink less when they do. Frequent drinking in young people is more strongly linked to dependence and harmful drinking subscale scores than quantity per occasion, yet it is the quantity per occasion that comprises more of the full AUDIT score in this group. Concurrently, frequency of consumption was not always a significant predictor of the AUDIT dependence subscale and a weak predictor of the harmful drinking subscale in older drinkers. This is a concern because we also found that the vast majority of an AUDIT score in this group comes from the question on frequency of consumption.

Research on young people “aging out” of their heavy episodic drinking (13-15) suggests that heavy episodic occasions are something that many younger drinkers can cease without intervention. In line with this, we were able to establish that the usual amount consumed per occasion is not a strong predictor of harmful or dependent drinking in young people, at least cross-sectionally. Please note that this is not to say that there aren’ t also young drinkers who will need help to decrease their consumption and could thus benefit from intervention. Rather, our results indicate that usual quantity per occasion is not a good predictor of these drinkers, while frequency of drinking may be. Longitudinal research to test this hypothesis could lead to work that better identifies those younger drinkers in need of intervention. This is important given the potential of early intervention to minimise problems in the longer term

Conversely, among older drinkers, in all but one of five-year cohorts aged 48 and over, usual quantity per occasion was a stronger predictor of both dependence and harmful drinking than frequency. Frequency of consumption has little predictive value for the dependence or harm subscales of the AUDIT in older drinkers. Indeed, in drinkers aged 73 and over the question on frequency of consumption was not significantly related to dependence. This is of particular concern because a very high proportion of AUDIT scores in older drinkers comes from the one question on frequency of consumption, as high as 87%. This number drops significantly when those who did not meet any cut-points of concern are removed from the sample. However, even then, nearly a third of the score on the AUDIT is coming from a question with very little predictive value for the rest of the scale. It may be that the question on frequency, that accounts for a high proportion of the total score while not being a strong predictor of the rest of the scale, is impeding more accurate identification of older drinkers who could benefit from intervention. A focus on quantity per occasion for this age group could be beneficial.

The primary limitation of the current study is the low response rates of the NDSHS. However, it is worth noting that the NDSHS’ sampling strategy is thorough and that their reported consumption levels, while lower than sales data, move with trends in sales (19).This study is also is based on self-report which is subject to biases such as social desirability (20). More specific to this study, the respondents who were missing on variables required for participation in this study appeared to drink less and experience less alcohol-related harm compared to those who remained in the sample, indicating that our sample may be heavier drinkers than would be found in the general population. Furthermore, the first two questions on usual quantity and frequency are asked with slightly different response categories than recommended in the AUDIT. As continuous measures of usual quantity and frequency are used in most of this paper, this will only have potentially had an impact on the data presented in Figure 4. However, replication of this work with AUDIT items as originally written is recommended. Finally, the AUDIT has been used in this study but questions on frequency of consumption in particular can be found in many screening tools such as the Alcohol, Smoking and Substance Involvement Screening Test (21) – work on how age based differences in drinking patterns influence the likelihood of being identified in other screening tools is also recommended.

Based on results from the current study, it does appear that clinicians and those screening for alcohol use disorders might wish to look out for young drinkers who are drinking like older people (frequently) and older drinkers who are drinking like young people (more per occasion). Unfortunately, as it currently stands, screening tools such as the AUDIT effectively do the opposite. Work replicating the findings here on longitudinal datasets would be worthwhile, followed by an age-specific investigation into how screening tools are scored and the potential for different scoring rubrics by age. Indeed, the authors would like to stress that we are not positing that questions on the usual quantity and frequency of consumption serve no purpose, but that it is instead the interpretation of responses to these questions that could be improved, particularly if the aim is to identify drinkers in need of intervention. Alternatively, a minimum score on the dependence or harmful drinking subscale could be used in conjunction with the AUDIT-C, as has been suggested elsewhere (e.g., 10).

## Data Availability

The Australian Institute of Health and Welfare manage the data collection and dissemination of the National Drug Strategy Household Survey and we are grateful to them for facilitating access to the data via the Australian Data Archive.

https://dataverse.ada.edu.au/dataverse.xhtml?alias=ndshs#:~:text=To%20apply%20for%20access%20to,sign%20the%20AIHW%20confidentiality%20undertaking.

## Acknowledgements

PMD is funded by an NHMRC Senior Research Fellowship (1136908) and has received investigator-driven funding from Gilead Sciences for work unrelated to this study. The Centre for Alcohol Policy Research is co-funded by the Foundation for Alcohol Research and Education, an independent, charitable organization working to prevent the harmful use of alcohol in Australia (**http://www.fare.org.au**).

## REFERENCES

1. GBD 2016 Alcohol Collaborators. Alcohol use and burden for 195 countries and territories, 1990–2016: a systematic analysis for the Global Burden of Disease Study 2016. The Lancet. 2018;392(10152):1015–35.

2. Whiteford HA, Degenhardt L, Rehm J, Baxter AJ, Ferrari AJ, Erskine HE, et al. Global burden of disease attributable to mental and substance use disorders: findings from the Global Burden of Disease Study 2010. The Lancet. 2013;382(9904):1575–86.

3. Department of Health. National Alcohol Strategy 2019-2028. Canberra: Australian Department of Health 2019.

4. Saunders J, Aasland O, Babor T, de la Fuente J, Grant M. Development of the Alcohol Use Disorders Identification Test (AUDIT): WHO Collaborative Project on Early Detection of Persons with Harmful Alcohol Consumption--II. Addiction. 1993;88(6):791 –804.

5. Babor T, Robaina K. The Alcohol Use Disorders Identification Test (AUDIT): A review of graded severity algorithms and national adaptations. International Journal of Alcohol and Drug Research. 2016;5(2):17–24.

6. Saunders J. AUDIT: Translations nd [Available from: https://auditscreen.org/translations.

7. Babor T, Higgins-Biddle J, Saunders J, Monteiro M. The Alcohol Use Disorders Identification Test: Guidelines for Use in Primary Care. 2 ed. Geneva: World Health Organization; 2001.

8. Donovan D, Kivlahan D, Doyle S, Longbaugh R, Greenfield S. Concurrent validty of the Alcohol Use Disorders Identification Test (AUDIT) and AUDIT zones in defining levels of severity among out-patients with alcohl dependence. Addiction. 2006;101:1696–704.

9. Lundin A, Hallgren M, Balliu N, Forsell Y. The use of Alcohol Use Disorders Identification Test (AUDIT) in detecting alcohol use disorder and risk drinking in the general population: validation of AUDIT using schedules for clinical assessment in neuropsychiatry. Alcoholism: Clincial and Experimental Research. 2015;39(1):158–65.

10. Selin KH. Alcohol Use Disorder Identification Test (AUDIT): what does it screen? Performance of the AUDIT against four different criteria in a Swedish population sample. Subst Use Misuse. 2006;41(14):1881–99.

11. Gmel G, Heeb J, Rehm J. Is frequency of drinking an indicator for problem drinking? A psychometric analysis of a version of the Alcohol Use Disorders Identification Test (AUDIT) in Switzerland Drug and Alcohol Dependence. 2001;64:151–63.

12. Australian Institute of Health and Welfare. National Drug Strategy Household Survey 2019. Drug statistics series no. 32. Cat. no. PHE 270. Canberra: AIHW; 2020.

13. Wakefield JC, Schmitz MF. How Many People have Alcohol Use Disorders? Using the Harmful Dysfunction Analysis to Reconcile Prevalence Estimates in Two Community Surveys.. Frontiers in Psychiatry. 2014;5.

14. Marmet S, Studer J, Bertholet N, Grazioli V, Daeppen J-B, Gmel G. Interpretation of DSM-5 alcohol use disorder criteria in self-report surveys may change with age. A longitudinal analysis of young Swiss men. Addiction Research & Theory. 2019;27(6):489–97.

15. Fillmore K, Midanik L. Chronicity of Drinking Problems among Men: A longitudinal study. Journal of Studies on Alcohol. 1984;45:228–36.

16. AIHW. National Drug Strategy Household Survey 2016: Detailed findings. Drug Statistics series no. 31. Cat. no. PHE 214. Canberra: AIHW; 2017.

17. O’ Brien H, Callinan S, Livingston M, Doyle JS, Dietze PM. Population patterns in Alcohol Use Disorders Identification Test (AUDIT) scores in the Australian population; 2007-2016. Australian and New Zealand journal of public health. 2020;44(6):462–7.

18. StataCorp. Stata/MP 15.0 for Windows. College Station TX 77845: StataCorp LP; 2017.

19. Livingston M, Callinan S, Raninen J, Pennay A, Dietze P. Alcohol consumption trends in Australia: Comparing surveys and sales-based measures. Drug and Alcohol Review [Internet]. 2018; 37(S1):[S9-S14 pp.].

20. Davis CG, Thake J, Vilhena N. Social desirability biases in self-reported alcohol consumption and harms. Addictive Behaviors. 2010;35(4):302–11.

21. Humeniuk R, Henry-Edwards S, Ali R, Poznyak V, Monteiro M. The Alcohol, Smoking and Substance Involvement Screening Test (ASSIST): manual for use in primary care.. Geneva: World Health Organization; 2010.

