## Supplementary Tables for "Age-based differences in quantity and frequency of consumption when screening for harmful alcohol use"

### Supplementary Table 1

### STROBE Checklist

| | Item No | Recommendation | Page No |
|---|---|---|---|
| **Title and abstract** | 1 | (*a*) Indicate the study's design with a commonly used term in the title or the abstract | 2 |
| | | (*b*) Provide in the abstract an informative and balanced summary of what was done and what was found | 2 |
| **Introduction** | | | |
| Background/rationale | 2 | Explain the scientific background and rationale for the investigation being reported | 3 |
| Objectives | 3 | State specific objectives, including any prespecified hypotheses | 3 |
| **Methods** | | | |
| Study design | 4 | Present key elements of study design early in the paper | 4 |
| Setting | 5 | Describe the setting, locations, and relevant dates, including periods of recruitment, exposure, follow-up, and data collection | 4 |
| Participants | 6 | (*a*) Give the eligibility criteria, and the sources and methods of selection of participants | 4 |
| Variables | 7 | Clearly define all outcomes, exposures, predictors, potential confounders, and effect modifiers. Give diagnostic criteria, if applicable | 4-5 |
| Data sources/ measurement | 8* | For each variable of interest, give sources of data and details of methods of assessment (measurement). Describe comparability of assessment methods if there is more than one group | *4-5* |
| Bias | 9 | Describe any efforts to address potential sources of bias | N/A |
| Study size | 10 | Explain how the study size was arrived at | 4 |
| Quantitative variables | 11 | Explain how quantitative variables were handled in the analyses. If applicable, describe which groupings were chosen and why | 4-5 |
| Statistical methods | 12 | (*a*) Describe all statistical methods, including those used to control for confounding | 5 |
| | | (*b*) Describe any methods used to examine subgroups and interactions | 5 |
| | | (*c*) Explain how missing data were addressed | 4 |
| | | (*d*) If applicable, describe analytical methods taking account of sampling strategy | N/A |
| | | (*e*) Describe any sensitivity analyses | 6 |
| **Results** | | | |
| Participants | 13* | (a) Report numbers of individuals at each stage of study—eg numbers potentially eligible, examined for eligibility, confirmed eligible, included in the study, completing follow-up, and analysed | 4 |
| | | (b) Give reasons for non-participation at each stage | 4 |

| | | | |
|---|---|---|---|
| | | (c) Consider use of a flow diagram | N/A |
| Descriptive data | 14* | (a) Give characteristics of study participants (eg demographic, clinical, social) and information on exposures and potential confounders | 4 |
| | | (b) Indicate number of participants with missing data for each variable of interest | 4 |
| Outcome data | 15* | Report numbers of outcome events or summary measures | 5-6 |
| Main results | 16 | (*a*) Give unadjusted estimates and, if applicable, confounder-adjusted estimates and their precision (eg, 95% confidence interval). Make clear which confounders were adjusted for and why they were included | 5-6 |
| | | (*b*) Report category boundaries when continuous variables were categorized | 4 |
| | | (*c*) If relevant, consider translating estimates of relative risk into absolute risk for a meaningful time period | N/A |
| Other analyses | 17 | Report other analyses done—eg analyses of subgroups and interactions, and sensitivity analyses | 6 |
| **Discussion** | | | |
| Key results | 18 | Summarise key results with reference to study objectives | 6-7 |
| Limitations | 19 | Discuss limitations of the study, taking into account sources of potential bias or imprecision. Discuss both direction and magnitude of any potential bias | 7 |
| Interpretation | 20 | Give a cautious overall interpretation of results considering objectives, limitations, multiplicity of analyses, results from similar studies, and other relevant evidence | 7 |
| Generalisability | 21 | Discuss the generalisability (external validity) of the study results | 7 |
| **Other information** | | | |
| Funding | 22 | Give the source of funding and the role of the funders for the present study and, if applicable, for the original study on which the present article is based | 5,8 |

**Supplementary Figure 1**

Diagram of exclusion and inclusion in analyses

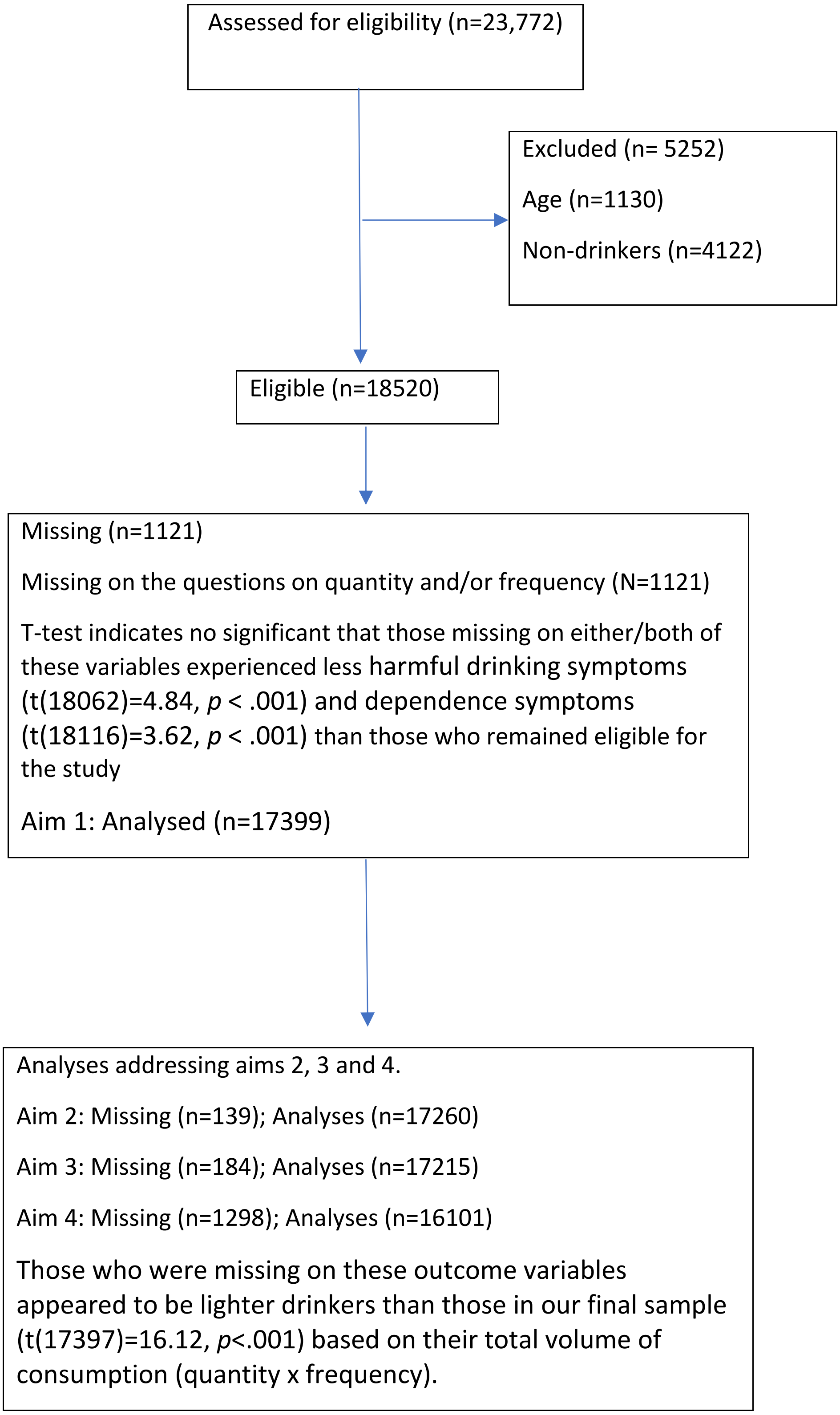

**Supplementary Table 2**

AUDIT questionnaire and equivalence in the National Drug Strategy Household Survey

| | AUDIT Item | NDSHS Item | Conversion |
|---|---|---|---|
| 1 | How often do you have a drink containing alcohol?<br>0. Less often<br>1. 1 day a month<br>2. 2-4 times per month<br>3. 2-3 times per week<br>4. 4 or more days per week | In the last 12 months, how often did you have an alcoholic drink of any kind?<br>1. Every day<br>2. 5 to 6 days a week<br>3. 3 to 4 days a week<br>4. 1 to 2 days a week<br>5. 2 to 3 days a month<br>6. About 1 day a month<br>7. Less often | The response options *every day* and *5-6 days a week* were coded into the *4 or more days per week* category of the AUDIT item, *1-2* and *3-4 days per week* were coded into the *2-3 times per week* category, *2-3 days a month* were coded as *2-4 times per month* and about *1 day a month* and *less often* were coded as *monthly or less*. |
| 2 | How many standard drinks do you have on a typical day when you are drinking?<br>0. 1-2<br>1. 3-4<br>2. 5-6<br>3. 7-8<br>4. 10 or more | On a day that you have an alcoholic drink, how many standard drinks do you have?<br>1. 20 or more drinks<br>2. 16-19 drinks<br>3. 13-15 drinks<br>4. 11-12 drinks<br>5. 9-10 drinks<br>6. 7-8 drinks<br>7. 5-6 drinks<br>8. 3-4 drinks<br>9. 1-2 drinks<br>10. Less than one drink | The response options of *9-10*, *11-12*, *13-15*, *16-19* and *20 or more* standard drinks were coded as *10 or more*, *7-8* was coded as *7-9*, *5-6* directly mapped onto its own category, as did the *3-4* drinks response. Finally, the *2 drinks*, *1 drink* and *half a drink* categories were all coded to the *1-2 drinks* category. |
| 3 | How often do you have six or more standard drinks on one occasion?<br>0. Never<br>1. Less than monthly<br>2. Monthly<br>3. Weekly<br>4. Daily or almost daily | Please record how often in the last 12 months you have had each of the following number of standard drinks in a day?<br>1. 20 or more standard drinks a day<br>2. 11-19 standard drinks a day<br>5. 7-10 standard drinks a day<br>7. 5-6 standard drinks a day<br>8. 3-4 standard drinks a day<br>9. 1-2 standard drinks a day<br>10. Less than 1 standard drink a day | The third item in the AUDIT is *"How often do you have six or more drinks on one occasion?"* To match the AUDIT as closely as possible responses were taken from a graduated frequency measure where the number of times an individual reported drinking 5-6, 7-10, 11-19 and 20 or more standard drinks were combined, and respondents were placed in the most appropriate category provided in the AUDIT categories. |
| 4 | How often during the last year have you found that you were not able to stop drinking once you had started?<br>0. Never | As written | N/A |

| | | | |
|---|---|---|---|
| | 1. Less than monthly<br>2. Monthly<br>3. Weekly<br>4. Daily or almost daily | | |
| 5 | How often during the last year have you failed to do what was normally expected of you because of drinking?<br>0. Never<br>1. Less than monthly<br>2. Monthly<br>3. Weekly<br>4. Daily or almost daily | As written | N/A |
| 6 | How often during the last year have you needed a first drink in the morning to get yourself going after a heavy drinking session?<br>0. Never<br>1. Less than monthly<br>2. Monthly<br>3. Weekly<br>4. Daily or almost daily | As written | N/A |
| 7 | How often during the last year have you had a feeling of guilt or remorse after drinking?<br>0. Never<br>1. Less than monthly<br>2. Monthly<br>3. Weekly<br>4. Daily or almost daily | As written | N/A |
| 8 | How often during the last year have you been unable to remember what happened the night before because you had been drinking<br>0. Never<br>1. Less than monthly<br>2. Monthly<br>3. Weekly<br>4. Daily or almost daily | As written | N/A |
| 9 | Have you or someone else been injured because of your drinking?<br>0. Never | As written | N/A |

| | | | |
|---|---|---|---|
| | 1. Less than monthly<br>2. Monthly<br>3. Weekly<br>4. Daily or almost daily | | |
| 10 | Has a relative, friend, doctor, or other health care worker been concerned about your drinking or suggested you cut down?<br>0. Never<br>1. Less than monthly<br>2. Monthly<br>3. Weekly<br>4. Daily or almost daily | As written | N/A |

**Supplementary Table 3**

Linear Regression models predicting dependence and harmful drinking subscales of the AUDIT with interactions between age and quantity and frequency respectively in order to establish the moderating role of age between quantity/frequency and subscale scores.

| | Quantity models | |
|---|---|---|
| | Dependence β | Harmful Drinking β |
| Age | -1.03*** | -2.31*** |
| Quantity | 5.11*** | 15.25*** |
| Age*Quantity | 0.09*** | 0.41*** |

| | Frequency Models | |
|---|---|---|
| | Dependence β | Harmful Drinking β |
| Age | -2.02*** | -0.77*** |
| Frequency | 77.06*** | 43.64*** |
| Age*Frequency | -0.80*** | -0.51*** |

*Please note that in the analysis using quantity, the average quantity per occasion
